# Network changes associated with right anterior temporal lobe atrophy: insight into unique symptoms

**DOI:** 10.1101/2025.01.29.25321213

**Authors:** Hulya Ulugut, Maria Luisa Mandelli, Anna Gilioli, Zoe Ezzes, Janhavi Pillai, David Baquirin, Amie Wallman-Jones, Amanda Gerenza, Eleanor R. Palser, Aaron Scheffler, Giovanni Battistella, Yann Cobigo, Howard J. Rosen, Zachary Miller, Kyan Younes, Bruce L Miller, Joel H. Kramer, William W. Seeley, Virginia E. Sturm, Katherine P. Rankin, Maria Luisa Gorno-Tempini

## Abstract

Semantic behavioral variant (also referred to as right temporal) frontotemporal dementia is a newly described syndrome associated with focal right anterior temporal lobe atrophy and a distinctive combination of behavioral and semantic changes. It is considered the right-sided counterpart of semantic variant primary progressive aphasia, with which it has overlapping neuropathological and cognitive mechanisms. Although more is known about how brain network alterations relate to both losses (e.g., word comprehension deficits) and gains (e.g., hyper-fluency) in cognitive and behavioral processes in the left-sided semantic progressive aphasia, less is known about these phenomena in the right-sided semantic behavioral variant. In this study, we investigated functional connectivity within the right counterparts of established ventral and dorsal cortical speech and language networks and their relationship to specific clinical manifestations in individuals with the semantic behavioral variant. We hypothesized that socioemotional-semantic deficits would be associated with reduced connectivity in the right ventral semantic network, while heightened behavioral manifestations, such as hyper-focus on specific interests (also referred to as rigidity), would be associated with increased connectivity in the right dorsal network. Using seed-based intrinsic connectivity analyses of fMRI data and cognitive scores from 22 individuals with semantic behavioral variant frontotemporal dementia and 48 cognitively normal individuals, we measured intrinsic connectivity strength in networks anchored in the right anterior middle temporal gyrus (ventral network) and in the right opercular inferior frontal gyrus (dorsal network). Functional connectivity values were then correlated with cognitive and behavioral measurements, controlling for global atrophy. Compared to the control group, individuals with semantic behavioral variant exhibited reduced connectivity in the ventral network (*t* = 2.7, *P* = 0.003), which was associated with socioemotional-semantic deficits (*r* = 0.47, *P* = 0.03), as measured by standardized tests. Conversely, increased functional connectivity was observed in the spared right dorsal network (*t* = 4.26, *P* < 0.001), which correlated with heightened scores for hyper-focus on fixed interests, as measured by retrospective ratings of medical notes (*r* = 0.63, *P* = 0.002). Together with previous evidence, these findings suggest that in individuals with focal anterior temporal lobe atrophy, enhanced behaviors such as hyper-focus and hyper-fluency are associated with altered functional dynamics within networks that remain relatively spared by the disease process. This highlights the complex interplay between damaged and spared networks in shaping the clinical manifestations of semantic behavioral variant frontotemporal dementia.

## Introduction

Frontotemporal dementia (FTD) is one of the leading causes of dementia before age 65 and most often manifests as behavioral problems (in behavioral variant FTD, [bvFTD])^1^ or speech and language impairment (in primary progressive aphasia, [PPA])^2^. These symptoms arise from focal neurodegeneration affecting either the frontal or temporal lobes, or both^1–3^. Behavioral problems in FTD are primarily characterized by interpersonal socioemotional deficits, which vary depending on the brain regions affected^1,4^. The term semantic behavioral variant frontotemporal dementia (sbvFTD) - also referred to as right temporal FTD^5–8^ or right semantic dementia (SD)^9–11^ - describes a clinical syndrome associated with predominant focal atrophy in the right anterior temporal lobe (rATL)^12^. Current cognitive neuroscience and clinical descriptions of sbvFTD indicate that the rATL serves as a semantic hub for socially and emotionally relevant information, such as knowledge of famous people, the meaning of facial expressions and paralinguistic cues, and other social concepts and norms^5,6,12–16^. Deficits in these semantic functions most often manifest clinically as loss of empathy^12^, with patients exhibiting difficulty understanding social situations and emotions, leading them to be unattuned or unresponsive to others’ needs and emotional states.

Growing evidence from studies in neurodegenerative diseases and functional MRI research in healthy individuals indicates that the ventral temporo-parietal network is the key anatomic substrate for lexical and object-related semantic processing. For instance, in the left hemisphere counterpart of sbvFTD, termed semantic variant PPA (svPPA), structural and functional connectivity changes are associated with deficits in word and object identification and are related to multimodal loss of semantic knowledge for objects^2,17–23^. Similarly, increasing evidence links right ventral network damage to semantic processing deficits for famous people and socioemotional information^12,13,24–28^. Disease-related degeneration of the ATL, a key hub in the temporo-parietal network, has been associated not only with cognitive and behavioral deficits but also with “heightened” behaviors, such as increased speech rate (referred to as hyper-fluency or press of speech) in individuals with svPPA^29^. However, individuals with sbvFTD with more right-lateralized degeneration show a hyper-focus on fixed interests^28^. This behavioral hyper-focus manifests across a spectrum of activities, from artistic pursuits to repetitive behaviors like clock-watching^5,6,9^. These symptoms have been regularly described over the years, but are heterogeneously labeled as rigidity, obsessions, compulsions, repetitive rituals, or preoccupations^5,7,9,12,30^, or more specifically as hyper-religiosity^31,32^ or hyper-graphia^32,33^. Recent retrospective work from an international group of experts reported that 78% of sbvFTD patients exhibit such behaviors at early stages of the disease^6^. Despite their high prevalence and relevance to everyday functioning, theoretical models focusing on the rATL do not fully explain the emergence of these behaviors in relation to structural atrophy. Some researchers suggest these behaviors stem from disturbances in reward processing, with a shift in hedonic valuation due to altered direct connectivity between the ATL and medial orbitofrontal regions^14,34^. Others attribute these behaviors to semantic loss, suggesting that deficits in comprehending multimodal (e.g., visual, auditory, chemosensory) socioemotional stimuli limit individuals’ options, leading to stronger personal preferences for stimuli that remain comprehensible to them^35–37^. While these hypotheses may account for the narrowness of interests, they do not explain why these activities become central to patients’ daily lives or why these actions are performed repeatedly with heightened motivation and extended attention spans (e.g., spending over eight hours each day writing^33^; see additional examples in Supplementary Table 1).

Although traditional lesion-based correlation studies have been instrumental in identifying symptoms associated with focal brain damage, such as linking ATL damage to semantic deficits in disorders affecting this region^13,38,39^, they have been insufficient for elucidating the neural mechanisms underlying clinical manifestations like hyper-focus on fixed interests (HFI). Studies using task-free functional MRI (tf-fMRI)^40^ have allowed the investigation of functional connectivity (FC) in focal neurodegenerative diseases^41–43^ with selective vulnerability to specific brain networks^44,45^. These studies provide evidence that degeneration in a critical network node can induce large-scale network-level changes^46,47^, potentially offering new insights into unique symptomatology. For instance, functional neuroimaging studies in svPPA revealed functional alterations and clinical correlations within left-hemisphere networks anchored in regions crucially involved in the disease^20,29^. Decreased FC in the ventral semantic network (seeding from the anterior portion of the middle temporal gyrus [aMTG] to the angular gyrus [AG]) correlated with verbal semantic deficits, the hallmark of svPPA. Conversely, increased FC in the left dorsal frontal network (seeding from the opercular part of the inferior frontal gyrus [opIFG] to the supramarginal gyrus [SMG]) was associated with an increased articulation rate^29^. These findings provide a compelling model for investigating altered functional responses to focal structural neurodegeneration in sbvFTD.

Substantial evidence links impairments in the right ventral network to socioemotional semantics, but research addressing the role of the right dorsal frontal network, anchored in opIFG, in individuals with neurodegeneration is limited. Resting-state FC studies in dementia suggest that decreased FC in this network is associated with attention deficits and apathy^48,49^. However, beyond the dementia literature, a large body of research in healthy individuals and those with neurodevelopmental disorders has indicated that increased FC in this network correlates with heightened task-related attentional load, action execution, rule acquisition, and enhanced responsiveness to positive outcomes^50–55^, which may explain symptoms such as hyper-focus on fixed interests (HFI).

In this study, we utilized seed-based tf-fMRI alongside clinical, cognitive and neuropsychological data from a cohort of individuals with early-stage sbvFTD and healthy controls (HC). We hypothesized that sbvFTD would cause FC alterations akin to those observed in svPPA within their ventral and dorsal right counterparts. Specifically, we hypothesized that the anatomical (right-side) counterpart of the ventral semantic network would show decreased FC, correlating with socioemotional semantic skills deficits, and that the counterpart of the frontal-dorsal network would show increased FC, correlating with enhanced HFI. This design aimed to provide new insights into the unique symptom combinations typical of sbvFTD.

## Materials and methods

### Participants selection

Patients with a clinical diagnosis of bvFTD^1^, svPPA^2^, SD^3^, and/or sbvFTD^12^ were identified from a database of individuals who participated in a prospective, multidisciplinary study on FTD at the Memory and Aging Center, University of California, San Francisco (MAC-UCSF). Patients with predominant rATL atrophy were identified using the atrophy lateralization index, as detailed in our previous publication^12^. Patients were excluded if: (i) they did not have a resting state fMRI sequence (n=30); (ii) their Mini-Mental State Examination (MMSE)^56^ scores were lower than 20 or clinical dementia rating (CDR)^57^ scores were equal or greater than 2 (n=2); (iii) structural MRI motion artifacts (n=5), white matter hyperintensities (Fazekas >2)^58^ (n=4), or fMRI artifacts (n=2) were present

Neurologically healthy, community-dwelling older adults (n=48) were recruited as a control group from a cohort enrolled in the BRain Aging Network for Cognitive Health study at the MAC-UCSF. Participants were verified as neurologically normal through a multidisciplinary assessment, which included neurological examination, neuropsychological and cognitive testing, and neuroimaging evaluation.

All participants, or their primary caregivers provided written informed consent for their participation in the study, and the experimental procedures were approved by the UCSF Human Research Protection Program.

### Functional, cognitive, and behavioral assessments

Each participant completed a comprehensive multidisciplinary evaluation that included neurological, functional, neuropsychological, socioemotional measures, as well as neuroimaging assessment^59^. The demographic, functional, cognitive, and behavioral profiles of the participants are summarized in Table 1. Descriptions of the assessment tools listed in Table 1 are provided in our previous publications^12,60^. Neurocognitive, behavioral, and disease severity data were collected within an average 90-day window before or after fMRI scanning.

**Table 1:** Sample Characteristics.

|  | sbvFTD (n=22) |  |  | HC (n=48) |  |  | P value |
| --- | --- | --- | --- | --- | --- | --- | --- |
|  | N | Mean | SD | N | Mean | SD |  |
| **Demographics** |  |  |  |  |  |  |  |
| Age (years) | 22 | 65.5 | 6.7 | 48 | 65.7 | 7.2 | N.S. |
| Sex (Female %)* | 22 | 50% | - | 48 | 50% | - | N.S. |
| Education | 22 | 16.6 | 3.0 | 48 | 17.5 | 2.0 | N.S. |
| Handedness (Righ/Left/AmbDx)* | 22 | 19/1/2 |  | 48 | 46/2/0 |  | N.S. |
| Scanner Type (Trio/Prisma) | 12/10 |  |  | 24/24 |  |  |  |
| **Functional Scales** |  |  |  |  |  |  |  |
| CDR | 22 | 0.8 | 0.4 | 48 | 0 | 0 | - |
| CDR-Box | 22 | 4.7 | 3.1 | 48 | 0 | 0 | - |
| FTLD Box | 22 | 6.7 | 3.7 | 48 | 0 | 0 | - |
| **Global Cognition** |  |  |  |  |  |  |  |
| MMSE | 22 | 26.1 | 2.9 | 38 | 29.3 | 0.8 | 0.00 |
| **Attention** |  |  |  |  |  |  |  |
| Digit Span Forward | 20 | 7.1 | 1.2 | 28 | 7.5 | 1.1 | N.S. |
| **Executive Functioning & Calculation** |  |  |  |  |  |  |  |
| Digit Span Backward | 19 | 5.1 | 1.0 | 38 | 5.8 | 1.4 | N.S. |
| Stroop (correct in 60 s) | 18 | 79.3 | 20.1 | 40 | 95.0 | 5.6 | N.S. |
| Design Fluency | 18 | 8.6 | 3.8 | 39 | 13.2 | 3.6 | 0.02 |
| Trials Time | 18 | 39.5 | 16.8 | 40 | 26.7 | 15.3 | 0.03 |
| Calculation | 19 | 4.5 | 0.6 | 31 | 4.7 | 0.5 | N.S. |
| **Rule Violation (RV)** |  |  |  |  |  |  |  |
| Verbal Fluency – Phonemic RV | 21 | 0.2 | 0.7 | 40 | 0.4 | 0.5 | N.S. |
| Verbal Fluency- Categorical RV | 21 | 0.06 | 0.2 | 40 | 0 | 0 | N.S. |
| Design Fluency RV | 18 | 0.3 | 0.6 | 40 | 0.1 | 0.3 | N.S. |
| **Visuospatial Processing** |  |  |  |  |  |  |  |
| Modified Rey–Osterrieth Figure - copy | 20 | 15.6 | 0.7 | 41 | 15.3 | 1.0 | N.S. |
| VOSP Number Location | 20 | 9.4 | 1.9 | 39 | 9.6 | 0.7 | N.S. |
| ***Episodic Memory*** |  |  |  |  |  |  |  |
| CVLT 30' short delay free recall | 20 | 4.9 | 2.7 | 6 | 8.8 | 0.4 | <0.001 |
| CVLT 10' long delay free recall | 20 | 3.5 | 2.9 | 6 | 8.4 | 0.5 | <0.001 |
| CVLT Recognition | 20 | 7.3 | 2.4 | 6 | 8.6 | 0.5 | <0.001 |
| Modified Rey–Osterrieth Recognition | 20 | 0.8 | 0.4 | 39 | 0.7 | 0.4 | N.S. |
| Modified Rey–Osterrieth- 10 min delay | 20 | 6.7 | 4.5 | 39 | 11.3 | 2.5 | 0.02 |
| ***Language*** |  |  |  |  |  |  |  |
| Verbal Agility | 19 | 5.4 | 0.9 | 17 | 5.8 | 0.3 | N.S. |
| Repetition | 19 | 2.9 | 0.2 | 39 | 3 | 0 | N.S. |
| WRAT Reading | 18 | 59.1 | 5.6 | 23 | 65.8 | 1.6 | 0.005 |
| Apraxia of Speech Rating | 10 | 0 | 0 | 9<sup>\$</sup> | 0<sup>\$</sup> | 0<sup>\$</sup> | N.S. |
| Dysarthria Rating | 10 | 0.1 | 0.3 | 59<sup>\$</sup> | 0.1<sup>\$</sup> | 0.07<sup>\$</sup> | N.S. |
| Syntax Comprehension | 19 | 4.5 | 0.7 | 16 | 4.9 | 0.2 | 0.02 |
| Lexical Fluency | 11 | 9.2 | 0.7 | 35<sup>\$</sup> | 14.9<sup>\$</sup> | 0.8<sup>\$</sup> | <0.001 |
| ***Verbal Semantics*** |  |  |  |  |  |  |  |
| Boston Naming Test | 19 | 8.4 | 3.6 | 40 | 14.5 | 0.7 | <0.001 |
| Verbal Fluency – Phonemic | 21 | 11.4 | 3.7 | 40 | 18.1 | 2.4 | <0.001 |
| Verbal Fluency - Categorical | 21 | 13.9 | 4.2 | 40 | 23.8 | 1.9 | <0.001 |
| Peabody Picture Vocabulary Test | 21 | 10.2 | 3.1 | 16 | 15.7 | 0.3 | <0.001 |
| ***Visual Semantics*** |  |  |  |  |  |  |  |
| Pyramids and Palm Trees Pictures % | 20 | 0.9 | 0.1 | 30 | 0.9 | 0.01 | <0.001 |
| ***Face Perception*** |  |  |  |  |  |  |  |
| CATS Face Matching | 20 | 11.5 | 0.8 | 15 | 11.8 | 0.3 | N.S. |
| ***Person Specific Knowledge*** |  |  |  |  |  |  |  |
| Famous Faces Naming % | 18 | 0.3 | 0.2 | 26<sup>\$</sup> | 0.7 | 0.2 | <0.001 |
| Famous Faces Familiarity % | 16 | 0.7 | 0.2 | 26<sup>\$</sup> | 0.8 | 0.1 | <0.001 |
| Famous Faces Semantic Association % | 17 | 0.7 | 0.2 | 26<sup>\$</sup> | 14.7 | 0.9 | <0.001 |
| ***Social Function & Emotion*** |  |  |  |  |  |  |  |
| CATS Affect Matching | 20 | 10.4 | 3.0 | 15 | 14.6 | 1.5 | <0.001 |
| Dynamic Affect Recognition Test | 8 | 6.7 | 1.9 | 71<sup>#</sup> | 10.1<sup>#</sup> | 2.1<sup>#</sup> | <0.001 |
| TASIT- Emotion Evaluation Test | 7 | 6.0 | 2.3 | 6 | 10.5 | 1.3 | <0.001 |
| TASIT- SI-M Sarcastic | 15 | 5.9 | 4.7 | 6 | 19.3 | 1.2 | <0.001 |
| TASIT- SI-M Sincere | 15 | 16.6 | 2.5 | 6 | 17.3 | 2.6 | N.S. |
| Cognitive Theory of Mind | 16 | 14.5 | 1.6 | 5 | 15.5 | 0.5 | N.S. |
| Emotional Theory of Mind | 10 | 12.3 | 1.3 | 5 | 15.7 | 0.3 | <0.001 |
| IRI- Empathic Concern | 14 | 20.3 | 8.6 | 18 | 27.8 | 5.2 | <0.001 |
| IRI- Perspective Taking | 14 | 14.3 | 6.0 | 18 | 24.1 | 7.0 | <0.001 |
| RSMS- Expressive Behavior | 16 | 5.1 | 3.0 | 5 | 22.6 | 5.1 | <0.001 |
| RSMS- Self Presentation | 16 | 10.3 | 5.1 | 5 | 26.4 | 5.4 | <0.001 |

\*= Categorical variables; <sup>#,\$</sup> = Since the present control group was not tested on these variables, we used data from Younes et al. (2022) (<sup>\$</sup>) and an independent group of 71 older healthy participants (<sup>#</sup>) to test differences in these scores between a control population and the sbvFTD patients enrolled in this study. CATS: Comprehensive Affect Testing System; IRI = Interpersonal Reactivity Index; PPVT = Peabody Picture Vocabulary Test; TASIT = The Awareness of Social Inference Test; RSMS = Revised Self-Monitoring Scale; SI-M = Social Inference- Minimal.
N represents the number of participants for which tests were available.

### Cognitive and behavioral scores for functional imaging correlations

For the brain-behavioral correlation analyses, we identified cognitive and behavioral measures hypothesized to be associated with the ventral or dorsal right networks in sbvFTD, based on previous literature. Specifically, we hypothesized that decreased connectivity in the right ventral network would correlate with reduced scores on tests assessing semantic processing of famous people and socioemotional concepts^13,16,24^. To quantify these constructs, we created composite scores from the UCSF Famous Face Battery (FFB)^25^, which includes confrontation naming, semantic association, and Familiarity Judgment tasks, and the Comprehensive Affect Testing System Affect Matching (CATS-AM) test^61^. Composite scores were calculated using the formula: ([test 1 patient’s total score/ test 1 maximum score) + (test 2 patient’s total score/ test 2 maximum score) + … (test n patient’s total score/ test n maximum score) / number of tests).

In previous studies on svPPA, increased connectivity in the left dorsal speech production network was linked to higher speech rate, suggesting that increased connectivity in spared networks may drive “positive” symptoms such as press of speech or hyper-fluency^29^. Drawing from the sbvFTD/right temporal literature, we proposed that hyper-focus on fixed interests, often termed rigidity or compulsive behaviors, might similarly be linked to increased dorsal connectivity^28,49,54,55^. Given the lack of a standardized assessment tool to quantify the presence or the severity of these symptoms, we devised a scoring protocol based on case notes.

Case notes were independently reviewed by four clinical researchers (HU, ZM, AG, AWJ) to assess the presence or absence of hyper-focus on fixed interests (HFI) symptoms. Following this initial assessment, reviewers collaborated by using a shared folder for discussion and resolving discrepancies through team consensus. In cases where symptoms related to HFI were present, the variety of the activities (Variety) and recurrence (Frequency) were further delineated. Total scores were calculated using the formula: ‘Variety’ x ‘Frequency’. The absence of the HFI symptom resulted in a score of 0. Variety ratings categorized activities as 1 for occurrence in a single domain (e.g. only cleaning), and 2 for involvement in multiple domains (more than one activity, e.g. cleaning, writing, religious practice). Frequency ratings classified activities as 1 for occasional occurrence (not part of daily routine, e.g. once a week), and 2 for activities integrated into the daily routine. Supplementary Table 1 presents examples from chart reviews and associated ratings.

### Brain MRI dataset

#### MRI acquisition

MRI images were acquired at the UCSF Neuroscience Imaging Center using two 3T Siemens scanners (Prisma or Trio). Magnetization-prepared rapid acquisition with gradient echo acquisition (MPRAGE) was used to acquire T1-weighted images. Parameters were identical for both scanners: 160 sagittal slices, isotropic voxel size of 1mm³, repetition time (TR) of 2300 ms, echo time (TE) of 2.98 ms, inversion time of 900 ms, flip angle of 9°, a field of view (FoV) of 256×256 mm², and an integrated parallel acquisition technique (iPAT) acceleration factor of 2. For fMRI, participants were instructed to lie still with closed eyes, ensuring they stayed awake. T2*-weighted volumes were acquired using an echo planar imaging (EPI) protocol. The Trio scanner captured 240 volumes of 36 AC/PC-aligned axial slices in an interleaved order, with TR/TE of 2000/27 ms, flip angle of 80°, slice thickness of 3 mm with a 0.6 mm gap, pixel size of 2.5×2.5 mm², field of view of 230×230 mm, and matrix of 92×92. The Prisma scanner captured 560 volumes of 66 slices with similar alignment, TR/TE of 850/32.8 ms, flip angle of 45°, slice thickness of 2.2 mm, pixel size of 2.2×2.2 mm², field of view of 211×211 mm², matrix of 96×96, and a multi-band acceleration factor of 6. Additionally, two volumes were acquired for distortion correction, using b=0 and opposite phase encoding directions (anterior/posterior and posterior/anterior).

#### Brain structural MRI pre-processing

Structural MRI data were processed using the Computational Anatomy Toolbox (CAT12) within the Statistical Parametric Mapping software framework (SPM12), running on Matlab 2021b. The data were enhanced using a spatial adaptive non-local means denoising algorithm^62^, followed by bias field correction, affine transformation for alignment, and processing using SPM’s “unified segmentation” protocol^63^. The images were segmented into gray matter, white matter, and cerebrospinal fluid, and then (i) spatially normalized to the Montreal Neurological Institute (MNI) reference space via an advanced geodesic shooting technique^64^, (ii) adjusted by the Jacobian determinants of the deformation field during spatial normalization to preserve tissue volume integrity, and (iii) output with a uniform isotropic voxel resolution of 1.5 × 1.5 × 1.5 mm³. Spatially normalized, segmented, and modulated gray matter images were finally smoothed using an 8 mm FWHM isotropic Gaussian kernel. Voxel-based inferential statistic was performed by fitting a general linear model in SPM12 on the smoothed and modulated GM tissue probability maps entering age, sex, handedness and total intracranial volume (TIV) as covariates of no interest. The statistical map showing gray matter volume differences between HC and sbvFTD was thresholded at P < 0.05 family wise error (FWE) corrected. The creation of the atrophy maps is further detailed in our previous publications^65,66^. The results of the atrophy maps are illustrated in Figure 1.

**Figure 1.**
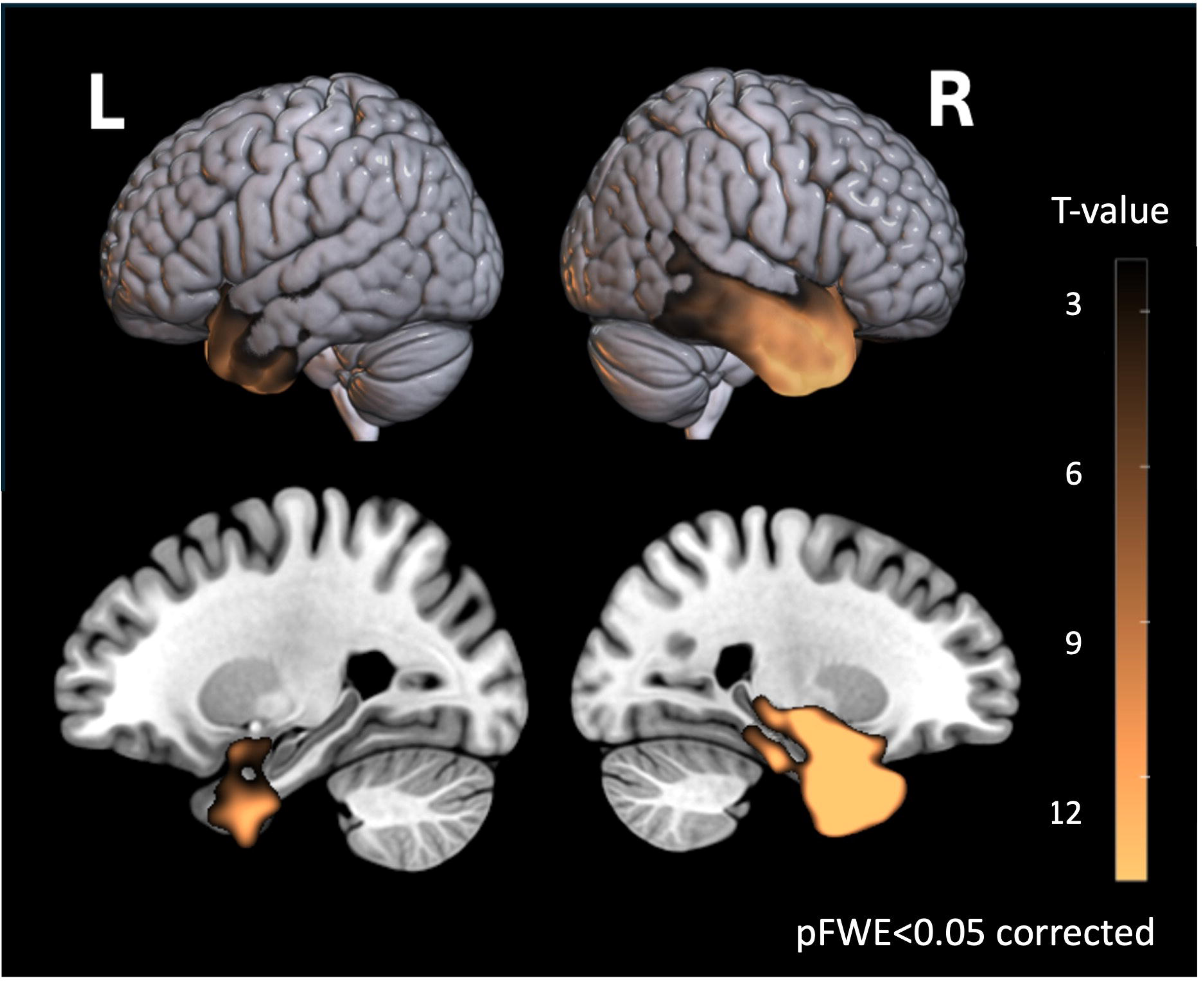

### Functional MRI data

#### Preprocessing of task-free fMRI

Functional MRI data were processed using a custom pipeline developed in Python, incorporating components from the Brain Software Library for Functional MRI (FSL), Statistical Parametric Mapping (SPM), Advanced Normalization Tools^67^ and the Analysis of Functional NeuroImaging (AFNI)^68,69^. To facilitate T1 equilibrium, the initial five volumes were excluded. The remaining volumes were slice-time corrected, realigned to the mean functional image, and assessed for rotational and translational head motion. EPI-related distortion artifacts were mitigated using FSL’s TOPUP utility, leveraging two acquired images with opposing phase-encode blip directions. The functional volumes were then registered to an EPI template in the MNI space using a combination of linear and non-linear warping. Spatial smoothing was applied using a 5-mm FWHM Gaussian kernel. Finally, the functional data were bandpass filtered within the frequency range of 0.008 Hz < f < 0.15 Hz, and non-signal fluctuations, including motion artifacts, their derivatives, and quadratic terms, along with white matter and cerebrospinal fluid signals, were regressed from the functional data as outlined in Satterthwaite and colleagues’ research^70^.

Subjects were included only if inter-frame head translations were less than 2 mm, head rotations were less than 2°, and motion spikes (defined as inter-frame head displacements greater than 1 mm) occurred in less than 5% of the total frames. Based on these criteria, 5 participants were excluded due to excessive motion or the presence of artifacts, and 2 participants were excluded due to white matter hyperintensity that could alter the interpretation of functional connectivity. The final cohort consisted of 22 individuals with rATL and 48 healthy controls.

#### Definition of seed-based functional connectivity analysis

The seed ROIs were defined as spheres of 5-mm radius centered at (1) the right anterior middle temporal gyrus (aMTG; MNI coordinates: *x* = 60, *y* = −6, *z* = −18) and (2) the pars opercularis of the inferior frontal gyrus (opIFG; MNI coordinates: *x* = 50, *y* = 8, *z* = 23) (Figure 1). Single-subject correlation maps were generated by calculating the *r*–Pearson correlation coefficient between the average blood-oxygen-level-dependent signal time course from each seed ROI and the time course from all other voxels of the brain. These correlation maps were converted to *z*-scores for parametric statistical comparisons.

The network anchored to the opIFG seed (right dorsal) included areas in the bilateral opercular and triangular part of the IFG, right middle frontal gyrus, right supramarginal gyrus, bilateral supplementary motor area and right inferior temporal gyrus (ITG). The network anchored to the aMTG seed (right ventral) included areas in the bilateral temporal pole, ITG, precuneus, hippocampus, and parahippocampal gyrus. Additional significant regions were located in the right AG, anterior insula, orbital IFG, bilateral anterior and posterior cingulate cortices, as well as in the bilateral orbital medial frontal gyrus, as previously described^19^.

#### Harmonization of Functional MRI data

To mitigate biases introduced by the use of different scanners and sequence parameters, we applied the ComBat method^71,72^ to the functional correlation maps after they were transformed into z-scores. This approach utilizes a multivariate linear mixed-effects regression that incorporates terms for both biological variables (age and sex) and scanner types to adjust for imaging feature measurements. Additionally, ComBat employs empirical Bayes techniques to refine the estimation of model parameters, which is particularly beneficial in studies with limited sample sizes. ComBat harmonization analyses were conducted using a publicly available R package, accessible at https://github.com/Jfortin1/ComBatHarmonization.

### Statistical Analyses

#### Demographic and clinical data

Descriptive statistics were calculated for all available demographic and clinical data. Between-group differences were assessed using t-tests for continuous variables and chi-square tests for categorical variables. All analyses were conducted using Python (version 3.10.9), with the significance level set at *P* = 0.05. Pearson correlation analyses were performed to assess the linear association between the variables of interest. To mitigate the potential impact of disease severity or global cognitive impairment, participants with a CDR ≥ 2 and/or an MMSE < 20 were excluded (see the participant selection section), thus such covariates were not included in the analyses.

#### Analysis of Functional MRI data

##### Group comparisons

Single-subject functional connectivity maps for each seed-based networks were entered into a voxel-wise two-sample *t*-test in SPM12 to identify functional connectivity alterations between the healthy cohort and the cohort of individuals with rATL. The cortical volume of the corresponding seed ROIs was included as covariates of no interest (age and sex are already modeled by ComBat). We included the total cortical volume as a measure of global atrophy. An implicit (binarized) mask of the networks of interest, defined at group level was used as previously defined in Battistella et al 2020^19^ (after FWE correction for multiple comparisons at peak level and k>180 for cluster extent in a healthy group of controls).

##### Brain-Behavior Correlations

We conducted Pearson correlation analyses with a one-sided test to examine specific directional hypotheses. We hypothesized that increased FC in the spared right dorsal network would be positively correlated with enhanced HFI scores, and that decreased FC in the damaged right ventral network would be positively correlated with lower scores on non-verbal socioemotional measures (FFB and CATS-AM). The one-sided test was chosen to align with these directional hypotheses, reflecting prior evidence and theoretical expectations about the relationship between FC and behavioral outcomes. While there was no missing data for the HFI, three participants had missing data for each of the FFB tasks. To ensure robustness, we performed the analyses both excluding these three participants and using an imputation method to address the missing values. The imputation analysis was conducted for sensitivity purposes. Detailed procedures for the imputation analysis are provided in supplementary material.

## Results

### Description of the cohort of patients

Table 1 summarizes the characteristics of the two cohorts included in the study. The healthy control group was demographically matched to the sbvFTD group; therefore, the two cohorts did not differ significantly in terms of sex, age, or education level. 18% of sbvFTD participants were either left-handed or ambidextrous. Of note, clinical symptoms were consistent with the anatomical distribution of cortical atrophy and the clinical presentation. All patients exhibited the same pattern of hemispheric lateralization as the right-handed participants. The average MMSE score was lower in the sbvFTD group (27 ± 2.7) compared to the HCs (29.8 ± 0.3), which is consistent with expectations based on published norms^56^. The mean CDR score was 0.8 (SD 0.4), indicating an early stage of the disease. Attention, calculation, and visuospatial functions were preserved, with no rule violations observed in verbal (phonemic/categorical) and design fluency tasks. Executive functions were generally within normal limits, except for mild to moderate impairments in the design fluency (*P* = 0.02) and the Trails tests (time, *P* = 0.03). As expected, sbvFTD patients exhibited severe impairments (*P* < 0.001) in visual semantic knowledge (measured by the Pyramids and Palm Trees pictures test) as well as verbal semantic knowledge (measured by the Boston Naming and Peabody Picture Vocabulary Tests). They also showed deficits in person-specific knowledge (measured by the Famous Face Battery [FFB]) (*P* < 0.001), although face perception skills remained intact. Affective knowledge was also impaired, as indicated by performance on the CATS-AM, Dynamic Affect Recognition Test, and the Awareness of Social Inference Test (TASIT) Emotion Evaluation Test (*P* < 0.001). Additional deficits were observed in lexical and verbal fluency, episodic memory, and recognizing paralinguistic cues (on the TASIT-sarcastic sections, while comprehension of sincere conversations was intact) (*P* < 0.001). Emotional perspective-taking was impaired (on the emotional Theory of Mind [ToM] test) (*P* < 0.001), although cognitive ToM was within normal limits. Informant-based surveys indicated diminished emotional sensitivity and reduced interpersonal reactivity, particularly in empathic concern and perspective-taking (*P* < 0.001). Lastly, the chart review rating of individuals with sbvFTD showed enhanced motivation and attention towards specific activities, many of which were part of their daily routine, often dedicating hours to these activities. Specific examples of this behavior are listed in Supplementary Table1, where it is defined as hyper-focus on fixed interests (HFI).

### Atrophy profile in sbvFTD

The distribution of atrophy exhibits the expected bilateral pattern within the temporal lobes, with the most significant peak observed in the right anterior temporal lobe (ATL) (MNI, x = 22, y = 3, z = −44; T-value = 22.22, cluster size = 108,072, *P* < 0.05, FWE-corrected). The atrophy pattern is most prominent in the inferior temporal gyrus (ITG), middle temporal gyrus (MTG), entorhinal cortex, and fusiform gyrus, extending to the orbital gyrus and amygdala (Figure 1).

### Functional connectivity

#### Altered functional connectivity in sbvFTD

Within the network seeded in the right opIFG, sbvFTD patients showed increased FC compared to HCs in a cluster localized in the right SMG (MNI coordinates of the cluster peak: 60, −32, 32; *T*-value: 4.26 cluster size: 98 voxels; *P* < 0.001 uncorrected) after controlling for global atrophy. No clusters with decreased FC were found in patients compared to controls within this network.

Within the network seeded in the right MTG, patients with sbvFTD showed decreased FC compared to HC individuals in a cluster localized in the right AG (MNI coordinates of the cluster peak: 46, −66, 34; *T*-value: 2.70 cluster size: 194 voxels; *P* = 0.003 uncorrected) after controlling for global atrophy.

#### Brain-Behavior Correlation in Altered Networks of sbvFTD

Correlation analyses conducted on clusters with significant FC alterations and the measures of interest revealed a significant correlation between higher HFI scores and increased FC in the right dorsal network (right SMG within the network seeded in the right opIFG) (*r* = 0.63, *P* = 0.001), assuming a positive directionality. Conversely, decreased FC in the right ventral network (between the right anterior MTG and the right AG) was associated with reduced socioemotional semantic skills (composite score of FFB and CATS-AM; *r* = 0.47; *P* = 0.03), assuming a positive directionality. The imputation analysis confirmed this positive association (beta = 0.12, SE = 0.053, *P* = 0.038). For visualization purposes, the imputed values of the missing participants were averaged over the total number of the imputed dataset (Figure 2).

**Figure 2.**
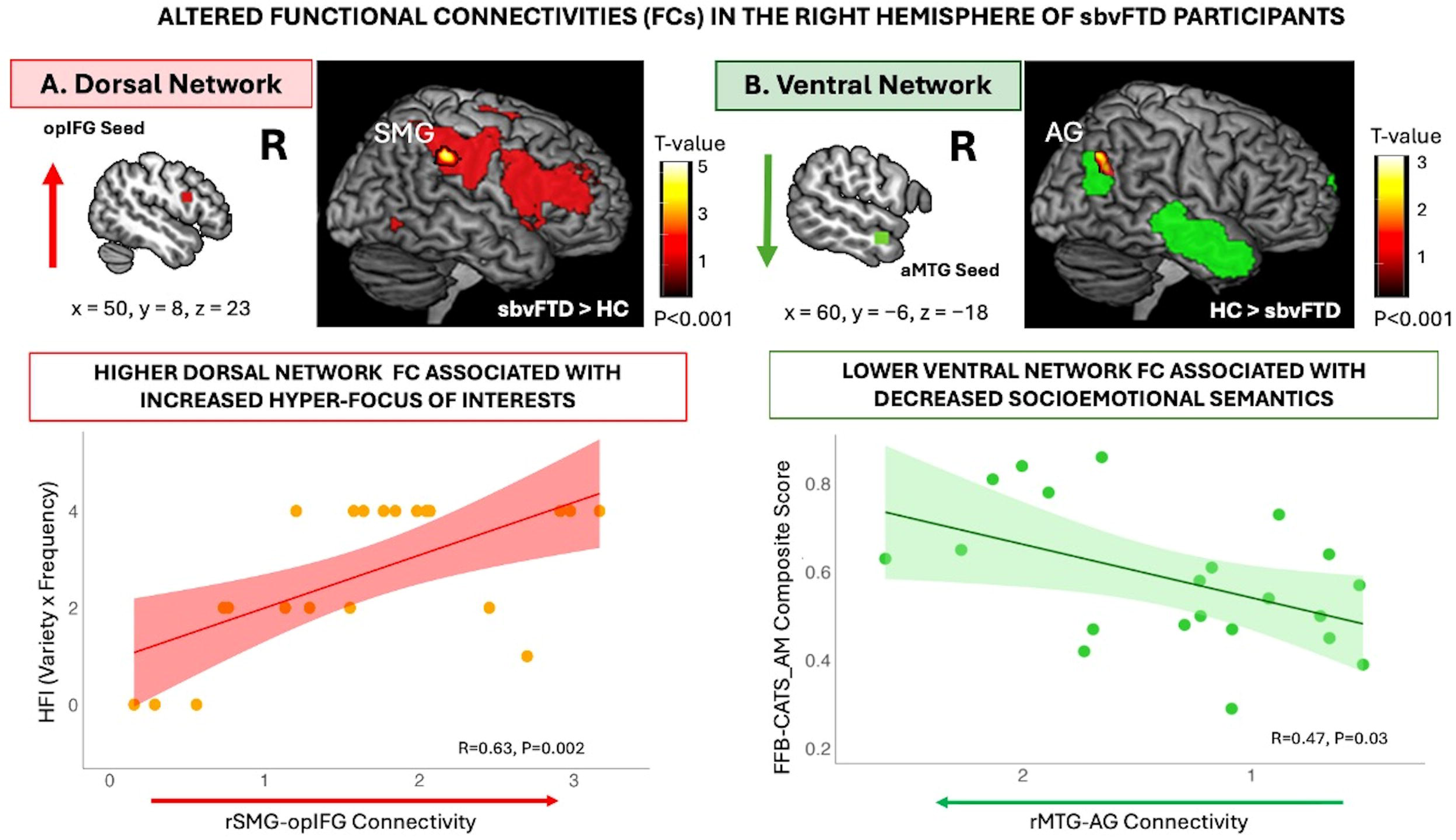

We also conducted a post-hoc analysis to further investigate the specificity of the correlation between the dorsal network and HFI scores. No significant correlations were observed between higher HFI scores and decreased FC in the right ventral network (*r* = 0.04, *P* = 0.75).

## Discussion

Neuroimaging and clinical data showed that increased functional connectivity within the right dorsal fronto-parietal network was linked to distinctive, rigid behavioral symptoms in patients with sbvFTD and focal anterior temporal lobe atrophy. In contrast, dysfunction within the right ventral temporo-parietal system was associated with deficits in socioemotional semantic processing. These results suggest distinct roles for each functional network in contributing to both impaired and heightened domains in sbvFTD. In the following paragraphs, we further examine the roles of these networks and cortical hubs in behavior and cognition.

Our findings indicate that both the dorsal and the ventral networks exhibited significant functional connectivity alterations with subregions of the inferior parietal lobule (IPL), specifically the supramarginal gyrus (connected to the inferior frontal seed) and the angular gyrus (connected to the middle temporal seed). Recent findings suggest that both the left and right IPL modulate connectivity within their respective subregions in a domain-specific manner^55^, with the left IPL consistently involved in language processing^19,20,73–75^. Here, we focus on the right IPL and its possible role in the specific “rigid” or “compulsive” behaviors described in sbvFTD, now described under the rubric of hyper-focus on fixed interest behavior^28^. Functional imaging, electrophysiological, and lesion-based studies have implicated the right IPL as integral to a system that facilitates flexible behavioral reconfiguration between two distinct operational modes^52,55,76^. Damage to the right IPL results in deficits related to (i) maintaining attentive control over current task goals and (ii) responding to salient new information or alerting stimuli in the environment. These impairments contribute to hemineglect, a classical syndrome often observed following lesions in this region^52^. Additionally, evidence from neurodevelopmental studies indicates that the right IPL selectively responds to both motoric and perceptual representations of actions, as well as socioemotional components of visual stimuli during action observation^50,51^. Previous parcellation studies in humans have delineated the right IPL and revealed distinct organizational patterns compared to nonhuman primate brains. These findings underscore the multidimensional role of the right IPL in cognition and behavior^77–80^. The forthcoming sections will elaborate on the specific engagement of each segment of the IPL within each intrinsic network.

### Right dorsal fronto-parietal network and its role in rigid, restricted behaviors

The lack of understanding of the neural mechanisms underlying this common symptom of hyper-focus on restricted, rigid interests in sbvFTD is a critical gap in the field^6,28^. From a behavioral perspective, previous studies propose that semantic loss and shifts in the hedonic system might underlie these factors^14,34,35,37^, but these theories were not empirically tested nor anatomically substantiated. One study found that individuals with semantic dementia (SD) used more efficient visual search strategies than healthy controls when the number of distractors increased^76^. The study revealed that conjunction search involving the highest number of distractors correlated with gray matter volume in the dorsal frontoparietal network, which was spared in the SD group. The researchers hypothesized that the enhanced attention skills observed in SD might arise from alterations in network connectivity due to decreased ventral stream information flow and enhanced function, but not altered structure, of gray matter in the dorsal frontoparietal network. However, their structural neuroimaging analyses did not allow them to directly show the FC alterations nor test hemispheric contributions^76^.

In the present study, we show increased FC in the right dorsal fronto-parietal network in sbvFTD and show that it correlates with symptoms attributable to hyper-focus on fixed interests. Previous work investigating the role of the IFG-SMG connectivity supports this hypothesis by showing its involvement in sustaining attention, detecting salient or novel events, phasic alerting, and task-set switching^52,81–84^, and highlighting its contributions to behavior^50,51,53–55,85–87^. Further functional segregation analyses identified five different IFG clusters related to their connections to other networks, specifically the opIFG and SMG, which are linked to task-specific attentional load and task execution^54^. Moreover, increased FC in this network has been attributed to heightened task-related activity and increased attentional load when targets and distractions are equal^88^, rule acquisition^89^, and activity modulation based on the outcome valence of choice tasks^90^.

Regarding the ‘impact of valence’, stronger activity increases were observed in a reward-seeking condition compared to a punishment-avoidance condition, indicating a stronger responsiveness to “better than expected” outcomes^90^. Finally, Uddin et al. (2021)^85^ underscored the role of this network in behavioral flexibility, while highlighting the contributions of other anatomical areas, such as the orbitofrontal cortex and dorsomedial striatum, especially in reward-related behavioral flexibility. Overall, these regions are crucial for maintaining focus on a task in the face of distraction and, conversely, for flexibly switching to new external demands when necessary for optimal behavior guidance^52^. In the case of sbvFTD, this system might lean towards “maintaining the task” rather than “switching to new stimuli”, which patients may have difficulty interpreting, especially if the system is overactive, as indicated in this study.

In this framework, structural damage and cognitive dysfunction in the right ventral semantic network in sbvFTD would shift interest away from activities involving nuanced, socially relevant knowledge, while relative structural sparing and increased connectivity in the dorsal circuit would increase attention to activities that require less interpersonal socioemotional engagement, such as collecting, writing, or exercising ^6,30,33^.

### Right ventral temporo-parietal network and its role in socioemotional semantics

We observed a correlation between lower FC in the aMTG-AG connection within the right ventral network and poorer performance on cognitive tests targeting specific semantic processes, such as famous face identification and emotion expression recognition. This finding indicates a role for this network in socioemotional semantics, relative to intact perception skills. A large body of functional neuroimaging literature depicts a major role for the AG in processing concepts rather than percepts when interfacing perception-to-recognition-to-action^21,91–93^. More specifically, the AG emerges as a cross-modal hub where converging multisensory information is combined and integrated to comprehend and give meaning to events, manipulate mental representations, solve familiar problems, and reorient attention to relevant information^94^. The pathway linking the aMTG and AG, connected by the middle longitudinal fasciculus ^95,96^, serves a distinct function in semantic processing^13,20,21,94,97^. This association has been studied on the left side^20^, supporting functions related to the control and manipulation of verbal and object-based semantics within the context of the large-scale default network. Specifically, individuals with svPPA showed decreased functional connectivity in the left ventral semantic network, correlated with weak word knowledge. While the right-sided network in sbvFTD has not been extensively studies, evidence consistently points to its hub regions being involved in socioemotional semantics as well^13,15,98^. Our findings contribute to the literature by confirming existing knowledge on ventral semantic networks and providing anatomical evidence for the clinical symptoms commonly seen in sbvFTD. This supports the use of more precise terminology that reflects the affected neural mechanisms.

It is noteworthy that the combination of damaged and spared neural networks, along with both decreased and increased connectivity, is necessary to produce such unique clinical manifestations. These manifestations likely occur in the early stage of the disease and eventually recede as it becomes more widespread. Such phenomena have also been described in the context of new talents emerging or changes in artistic expression in focal neurodegenerative diseases^76,98–100^. Future studies employing novel methodologies are warranted to elucidate how neural dynamics alter in response to focal neurodegeneration over the course of the disease, leading to changes in clinical manifestations over time.

### Limitations

The study has several limitations. First, our sample size was relatively small (n=22), which may limit the generalizability of our results. However, this is a rare disease, and it is nevertheless a large cohort compared to other published reports. Additionally, we created a highly specific patient cohort by excluding individuals with severe global cognitive decline and higher disease severity, focusing on those with focal atrophy in the temporal lobe, as measured using an objective atrophy index. All patients included in this analysis were recruited for a prospective, multidisciplinary, longitudinal observational study. Some data were missing for certain tests, often due to updates in administered test batteries over time or patient fatigue. We addressed this by imputing missing data using well-established relationships, particularly between tests for famous people knowledge and atrophy of the right temporal lobe, as well as estimating based on detailed notes recorded during the tests. Lastly, because there are no previously validated objective tests for measuring engagement in these hyper-focused behaviors, we had to rely on chart reviews. To mitigate potential biases, four independent raters reviewed the case notes, providing assessments of the frequency and variety of HFI behavior. While not standardized, this approach was a necessary adaptation that allowed for the inclusion of this important behavioral measure in our analysis.

## Conclusion

This study provides novel neural insights into the poorly understood symptoms associated with focal right anterior temporal lobe degeneration. Our findings uncover alterations of both increased and decreased network connectivity in sbvFTD, extending beyond traditional atrophy-based approaches. These results deepen our understanding of the neurobiological basis of clinical manifestations in focal neurodegeneration.

## Data availability

Upon journal publication, individual-level data from this study will be shared in an access-controlled FAIR Alzheimer’s Disease Data Initiative AD Workbench repository at https://fair.addi.ad-datainitiative.org.

## Acknowledgements

The authors would like to sincerely thank the participants and their families for their invaluable time, effort, and commitment to the research. Their dedication and cooperation were essential to the success of this study, and we deeply appreciate their contributions to advancing our understanding of this important area of research.

