## Supplementary Table 1 for "Network changes associated with right anterior temporal lobe atrophy: insight into unique symptoms"

| **Cases** | **Symptom** | **Score (VxF)** | **Real life examples from chart reviews** | **Range of activities** | **Variety (V)** | **Recurrence of activities** | | **Frequency (F)** |
| --- | --- | --- | --- | --- | --- | --- | --- | --- |
| Case 1 | Present | 4 | *…spends his time for checking the calendar and lining up clocks so that they read the same time at home… he will stir his coffee 60 times and will take his pills only at a certain time daily… hoarding objects during a move to Kansas… he wanted to journal everything…* | multiple | 2 | Part of daily routine | | 2 |
| Case 2 | Present | 2 | *… becoming more regimented with his routine, being especially content to stay home every day, clean the house, and do laundry according to a regular schedule...* | only cleaning | 1 | Part of daily routine | | 2 |
| Case 3 | Present | 4 | *… becoming very concerned about a healthy diet and started to have a regimented eating schedule and diet… developed new interests in word jumbles, solitaire, and dominos. She would play them at a specific time of the day. Her husband reports that the patient has lived by her strict schedules for the past couple of years. She schedules most daily activities, including regimented eating, exercise, taking medications, or playing games.* | multiple | 2 | Part of daily routine | | 2 |
| Case 4 | Present | 4 | *… developed some compulsive habits… needs to watch Jeopardy at 7:00 PM every night regardless of what else is happening in the house. He has also become obsessed with number games such as sudoku or similar games. He has purchased many books with sudoku puzzles and spends a significant amount of time playing these types of games on the internet. He sometimes has ignored other things in the house in order to do this. He has also developed a habit of wanting to light a fire in the fireplace nearly every night, and he has a compulsive tendency to prod at the fire with the poker very frequently...* | multiple | 2 | Part of daily routine | | 2 |
| Case 5 | Present | 4 | *… the patient has become significantly more rigid in her ways - she had always been a healthy eater but become pickier for her daily diet over the past few years… has developed newfound obsessions about retirement and her financial security to the point where it inappropriately becomes the central topic of her conversations…* | multiple | 2 | Part of daily routine | | 2 |
| Case 6 | Absent | 0 | 0 | 0 | 0 | 0 |  |  |
| Case 7 | Present | 2 | *… fixed in time and punctuality/She is extremely focused on good health and says she is "determined to live a long life."  She eats an extremely specific breakfast every day with multiple ingredients that she knows the exact nutritional value of…* | health | 1 | Part of daily routine | | 2 |
| Case 8 | Present | 4 | *… became more rigid in daily activities, excessively documenting his laboratory tests on an electronic tablet… became more rigid in how he wanted his shirts folded by his wife, not allowing creases on the sleeves, even though she had been folding them in a specific manner for years…* | multiple | 2 | Part of daily routine | | 2 |
| Case 9 | Absent | 0 | 0 | 0 | 0 | 0 |  | 0 |
| Case 10 | Present | 4 | *… developed new interests that bordered on an obsessional quality (sport teams, wearing team gear while watching games)/ his life has become more regimented and rigid: eat a banana every morning, he has specific chores he performs on specific days, wear only the color blue…* | multiple | 2 | Part of daily routine | | 2 |
| Case 11 | Present | 1 | *…he can perseverate a lot such as over the fact that they would have to pay here today despite being told repeatedly by her that this wasn't the case, but no other examples of obsessions and compulsions. He has also become rigid in his weekly habits, and is adamant about going to the movies every Saturday…* | only movies | 1 | occasionally | | 1 |
| Case 12 | Present | 2 | *…had obsessions with a metal detector looking for metal objects on his property, also the website ancestory.com... but the caregiver reports that the obsessions are not often…* | multiple | 2 | occasionally | | 1 |
| Case 13 | Present | 4 | *…"compulsive hoarding became an issue" in their move and her son recalls her keeping too many objects some called "orphan objects" from thrift stores…eats same things every day… fixation on scratching the scab off the back of her hand so it can not properly heal… also fixated on the "tissue fat" of her thighs and wipes her nostrils frequently saying they are dripping but is unclear whether she actually has a runny nose or not… repetitive behavior of "humming/mumbling sound" when sitting idly or walking.* | multiple | 2 | Part of daily routine | | 2 |
| Case 14 | Present | 2 | *…won’t walk under powerlines, same route whenever she walks and "obsession with political and charitable organizations" … also enjoys sudoku and crossword puzzles, although it is not a daily activity…* | multiple | 2 | occasionally | | 1 |
| Case 15 | Present | 4 | *…obsessed with making kids happier... she began to buy excessive amounts (full suitcases) of cheap toys… she would obsessively monitor her partner how he was caring for his children, such as how to set up the car seat, or how to cut up their food... had a preoccupation with young men, talking to them in public and saying how nice they looked, and printing pictures of young men… She is not rude, but clearly makes them uncomfortable with her attention... She encourages young men to hug her... She has also begun to display inappropriate attention to young children she doesn’t know, touching them… She does like to do variety puzzles and words with friends, and will spend five hours per day doing these…* | multiple | 2 | Part of daily routine | | 2 |
| Case 16 | Present | 4 | *…Became rigid with his eating regime… only eating popcorn, pizza and cookies… He eats popcorn at almost every meal of the day. When they buy a pizza on a given day of the week, he eats only one slice that day and thereafter eats one slice per day for the rest of the week until the pizza is finished... He also carries rolls of mint tabs in his pocket and eats them repeatedly during the day and even at night sometimes…* | multiple | 2 | Part of daily routine | | 2 |
| Case 17 | Present | 4 | *…She received mail solicitations from the Publisher’s Clearing House and political groups, and she sent money to these groups. She accumulated so many subscriptions and gifts from the Publisher’s Clearing House… Spends a lot of time rowing… developed new dietary preferences… She became interested in non-GMO, “pure” foods after reading a book titled “Eat Naked”… She has continued this “pure” diet since - 2014 - obsession with John Travolta, Tom Cruise and defends scientology…* | multiple | 2 | Part of daily routine | | 2 |
| Case 18 | Present | 2 | …saves toilet paper and delays flushing toilet to save water. Refuses to shower because it makes her itchy… | only saving water | 1 | Part of daily routine | | 2 |
| Case 19 | Present | 4 | *…developed a heightened interest for Facebook and amazon in the past 3-4 years, often sharing religious “blurbs” and relatively personal information about his family’s life... Also buying pens and watches (over 100)…* | multiple | 2 | Part of daily routine | | 2 |
| Case 20 | Present | 2 | *…became accustomed to daily routines in which she would do nothing more than spending the whole day driving to Target or Starbucks multiple times, and walking around everywhere when she was no longer able to drive after a car accident ...* | only driving/ walking to certain location | 1 | Part of daily routine | | 2 |
| Case 21 | Absent | 0 |  |  | 0 |  |  | 0 |
| Case 22 | Present | 4 | *… new interest in paranormal podcasts, increased need to make lists, and her desire not to throw food away, sometimes eating items her husband says are no longer good (she does know she should not pick up food off the trash)…* | multiple | 2 | Part of daily routine | | 2 |
