## Supplementary Material for "Network changes associated with right anterior temporal lobe atrophy: insight into unique symptoms"

**Missing data handling**

Three patients had documented clinical descriptions of impairment severity for the famous faces battery (FFB) but lacked exact scores for all of the three tasks. To estimate these missing scores, we applied Multiple Imputation by Chained Equations (MICE)^1^ using the mice package in R. Prior evidence links bilateral temporal lobe involvement in face processing, with right temporal regions associated with recognition and left temporal regions with naming^2–4^. Our data support this association, showing positive correlations between the FFB naming component and atrophy in both the left (R = 0.56, p = 0.007) and right (R = 0.66, p = 0.001) temporal lobes. However, the triplet (R = 0.42, p = 0.046) and recognition (R = 0.50, p = 0.025) components correlated only with the right temporal atrophy. Consequently, the total FFB score correlated only with right temporal atrophy (R = 0.65, p = 0.002), which was used as the predictor for imputing the FFB total score. Predictive Mean Matching was employed to ensure that each imputed FFB value closely matched the distribution of observed values. For each imputation iteration, the composite score was calculated based on imputed FFB values, and the distribution of imputed data was inspected to ensure alignment with observed data. Twenty imputed datasets were generated, with simple regression models estimated and pooled according to Rubin’s rules^5^.
